# Measuring brain integrity using MRI: a novel biomarker for Alzheimer’s disease using T2 relaxometry

**DOI:** 10.1101/2020.05.13.20100339

**Authors:** Alfie R. Wearn, Volkan Nurdal, Esther Saunders-Jennings, Michael J. Knight, Hanna K. Isotalus, Serena Dillon, Demitra Tsivos, Risto A. Kauppinen, Elizabeth J. Coulthard

## Abstract

Early Alzheimer’s disease diagnosis is vital for development of disease-modifying therapies. Prior to significant loss of brain tissue, several microstructural changes take place as a result of Alzheimer’s pathology. These include deposition of amyloid, tau and iron, as well as altered water homeostasis in tissue and some cell death. T2 relaxation time, a quantitative MRI measure, is sensitive to these changes and may be a useful non-invasive, early marker of tissue integrity which could predict conversion to dementia. The different factors that affect T2 may cause it to increase, as in the case of free water, or decrease, as in the case of iron, amyloid and tau. Thus, tissue affected by early Alzheimer’s disease could become more heterogeneous yet show no change in average T2. We hypothesise that T2 heterogeneity in regions affected early in Alzheimer’s disease (hippocampus and thalamus) may present a sensitive early marker of microstructural changes in Alzheimer’s disease.

In this cohort study, we tested 97 healthy older controls, 49 people with mild cognitive impairment (MCI) and 10 with a diagnosis of Alzheimer’s disease. All participants underwent structural MRI including multi-echo sequence for assessing quantitative T2. Cognitive change over one year was assessed in participants with MCI. Hippocampus and thalamus were automatically masked using ASHS and Freesurfer, respectively. T2 distributions were modelled using log-logistic distribution giving measures of log-median value (midpoint; T2μ) and distribution width (heterogeneity; T2σ).

We show an increase in heterogeneity (T2σ; p<.0001) in MCI compared to healthy controls, which was not seen with midpoint (T2μ; p=.149) in the hippocampus and thalamus. Hippocampal T2 heterogeneity predicted cognitive decline over one year in MCI participants (p=.018), but midpoint (p=.132) and volume (p=.315) did not. Age affects T2, but the effects described here are significant even after correcting for age.

We show that T2 heterogeneity can identify subtle changes in microstructural integrity of brain tissue in prodromal Alzheimer’s disease. We describe a new model that takes into account the competing effects of factors that both increase and decrease T2. These two opposing forces act in opposition and mean that previous human literature focusing on midpoint T2 has obscured the true potential of T2 as an early marker of Alzheimer’s disease. In fact, T2 heterogeneity outperforms midpoint and volumetry in predicting cognitive decline in those with MCI. We propose that T2 heterogeneity reflects microstructural integrity with potential to be a widely used early biomarker of Alzheimer’s disease.

## Introduction

Alzheimer’s disease treatments and therapies that stop or slow down neuropathology will be most effective if administered as early as possible, before significant neurodegeneration has occurred. Accurate early Alzheimer’s disease diagnosis is vital to identify appropriate clinical trial study groups of “at-risk” individuals to expedite development of new compounds (Cummings *et al*., 2014; Alzheimer’s Association, 2015) and to target disease-modifying treatments when available.

Structural and quantitative MRI show promise in their ability to identify changes in the brain that indicate early Alzheimer’s pathology. Measuring the volume of the hippocampus and entorhinal cortex has been shown to predict progression of mild cognitive impairment (MCI) to Alzheimer’s disease (Fox *et al*., 1999;Jack *et al*., 1999; Jack *et al*., 2000; Jack *et al*., 2003;Jack *et al*., 2004; Fleisher *et al*., 2008;Henneman *et al*., 2009;Teipel *et al*., 2013). Detectable change in volume is indicative of significant tissue loss,which is likely to be irreversible. As treatment with disease-modifying therapies would be optimal before such significant macrostructural change, we ask whether MRI could be used to identify microstructural changes that occur earlier in the disease-course, before significant volume loss.

Prior to significant loss of tissue volume, several microstructural changes take place as a result of Alzheimer’s disease pathology – i) oligomers and plaques of β-amyloid (Aβ) and neurofibrillary tangles (NFTs) build up around the MTL and the thalamus (Braak and Braak, 1991;Braak and Braak, 1995;Selkoe and Hardy, 2016), ii) iron is elevated in the brains of people with MCI and Alzheimer’s disease (Smith *et al*., 2010) and iii) even small amounts of necrosis leading to breakdown of cell membranes and oedema, will increase the motility of water within a given region. Increase in water motility is not necessarily specific to Alzheimer’s disease, and can occur in healthy ageing (Golomb *et al*., 1993; Pol *et al*., 2006; Frisoni *et al*., 2008). Accurately measuring such microstructural changes may allow identification of tissue that is at risk of degradation or has reduced functionality compared to a previous state.

T2 relaxometry is an MRI approach that may be able to report microstructural tissue integrity. Relaxation time is a measure, detectible by MRI, that describes the time taken for protons to return to a state of equilibrium following electromagnetic excitation. Specifically, T2 relaxation describes the transverse component of magnetisation. T2 relaxation time of biological tissue varies depending on its physical properties and its surrounding environment. It is primarily driven by water content and mobility and the presence of macromolecular structures and paramagnetic materials e.g. iron (Symms *et al*., 2004). For example, pure water will have a very long relaxation time, whereas in fatty substances T2 will decay much quicker. T2 is therefore sensitive to microscopic and physico-chemical tissue properties that can change as a result of pathology.

Given that quantitative T2 can be easily measured on routine MRI scans, adding just a couple of minutes to standard T2-weighted structural scanning times, it has been previously explored as an early marker of Alzheimer’s disease pathology. However, previous studies on the effect of Alzheimer’s pathology on T2 in the human brain have yielded varied and sometimes contradictory results. Most studies describe a prolonged T2 in the hippocampus of those with Alzheimer’s disease (Kirsch *et al*., 1992; Laakso *et al*., 1996; Pitkanen *et al*., 1997; Wang *et al*., 2004; Dawe *et al*., 2014), whereas others find the opposite (House *et al*., 2006;Luo *et al*., 2013) or no change at all (Campeau *et al*., 1997) (see Tang *et al*. (2018) for a comprehensive review). In two studies (Kirsch *et al*., 1992;Laakso *et al*., 1996), change in thalamic T2 was not associated with Alzheimer’s pathology or cognitive impairment, but T2 increased with age (Laakso *et al*., 1996). In another study,Dawe *et al*. (2014) found that Alzheimer’s pathology was associated with decreased T2 within the thalamus.

These inconsistencies in human literature are not fully reflected in studies of transgenic rodent models of Alzheimer’s disease, which consistently show a decrease in hippocampal T2 (Tang *et al*., 2018). Inconsistencies within the human literature, and between human and animal studies could be a consequence of the multiple pathological processes occurring in the human brain that have opposing effects; either shortening or lengthening T2. In contrast, mouse models are usually dominated by a single pathological process such as amyloid deposition.

Increased water content, such as that caused by increased CSF, oedema or cell membrane damage, will prolong T2 (Laakso *et al*., 1996). Conversely, increase in iron (Meadowcroft *et al*., 2015) or in macromolecule-to-water ratio due to accumulation of high density protein aggregates, such as Aβ, shorten T2 (House *et al*., 2008). Even in early stages of the pathological progression of Alzheimer’s disease within the brain, factors which cause T2 to either increase or decrease are both occurring in early-affected regions such as the hippocampus and the thalamus (Braak and Braak, 1991; Braak *et al*., 1996; Thal *et al*., 2002; Ward *et al*., 2014). Either effect may be more or less dominant in clusters throughout these regions. Averaging across the entire region could therefore yield, on average, a net change in T2 of zero. A change in the average value of T2 would only come about if T2-shortening factors dominate over T2-prolonging factors or vice-versa, which may not be the case in the earliest stages of the disease. Equal dominance relative to each other would cause a net change in the mean value of zero. We, therefore, aim to assess the use of the width of the distribution of T2 as a marker of tissue heterogeneity and microstructural integrity. We hypothesise that T2 distribution width will be wider in more heterogeneous tissue present in the early stages of Alzheimer’s pathology (at the stage of MCI), and will be more sensitive to early pathology than a midpoint measure of the T2 distribution.

T2 heterogeneity as a marker of tissue integrity is a novel measure, with only two known previous studies of its utility. One of these demonstrates that T2 heterogeneity is a useful measure in accurately determining stroke onset time in an animal model (Norton *et al*., 2017). The other presented pilot data from our group, concluding that T2 heterogeneity can improve accuracy in distinguishing between healthy controls, those with MCI and Alzheimer’s disease patients (Knight *et al*., 2019). Here we expand this work by describing a model of T2 dynamics through the course of Alzheimer’s disease, in comparison to healthy ageing, with a view to creating a practical biomarker which may identify Alzheimer’s disease neuropathology prior to significant tissue atrophy. We also assess the ability of hippocampal T2 heterogeneity (as opposed to T2 midpoint or volume) to predict cognitive decline in people with MCI.

## Materials and Methods

The analyses in this paper combine data from two longitudinal studies similar in cohort demographics and study design. No participants took part in both studies. Both studies are detailed in the following section. Where data collected are not identical between cohorts, we have normalised equivalent metrics within cohort and combined data after normalisation.

### Participants

Participants fulfilling the Petersen criteria (Albert *et al*., 2011) for diagnosis of MCI were recruited to both studies (Study 1: n=30, Study 2: n=29). Healthy older people (HC), with no history of memory problems or significant neurological disorders were recruited as controls to each study (Study 1: n=61; Study 2: n=56). All healthy controls had Montreal Cognitive Assessment (MoCA) > 26 (study 1) or Addenbrookes Cognitive Examination 3 (ACE-III) > 88 (study 2). 7 participants originally recruited as healthy controls in study 1 were found to have MoCA scores of <26, so were reclassified as MCI. Study 1 also included patients with diagnoses of Alzheimer’s disease (AD) who retained capacity to consent (n=10). These sample sizes are in-line with similar studies on brain structure abnormalities in MCI and Alzheimer’s disease, and are sufficient to observe significant differences in hippocampal volume.

Subjects for both studies were recruited from local GP surgeries and memory clinics in the Bristol area (having received MCI diagnoses or reported memory problems), Join Dementia Research, Avon and Wiltshire Mental Health Partnership’s Everyone Included system, an in-house database of volunteers, replies to poster adverts or through word of mouth. All patients provided informed written consent prior to testing as according to the Declaration of Helsinki. Ethical approval was given by Frenchay NHS Research Ethics Committee.

The current analyses included all participants who had volumetry and T2 relaxometry data for both hippocampal subfields and thalamus, study 1 n=90 (50 HC, 30 MCI, 10 AD), study 2 n=66 (47 HC, 19 MCI). See tables 1 and 2 for demographic details.

A total of 10 MCI participants were followed-up after one-year from each study. Cognitive function was tested at baseline and follow-up using the MoCA in study 1 and the ACE-III in study 2.

### Imaging parameters

Scans for both studies were acquired at CRICBristol, University of Bristol, UK, on the same Siemens Magnetom Skyra 3T system equipped with a parallel transmit body coil and a 32-channel head receiver array coil. The two studies used similar, but slightly different scanning protocols.

### Study 1

This protocol has been previously described by Knight *et al*. (2019). The imaging protocol included a 3D T1-weighted whole-brain magnetization prepared rapid acquisition gradient-echo (MPRAGE) and 2D multi-contrast multi-spin-echo (CPMG).

MPRAGE: Coronal, whole-brain, repetition time (TR) 2200 ms, Echo Time (TE) 2.42 ms, Inversion time (TI) 900 ms, flip angle 9°, acquired resolution 0.68 x 0.68 x 1.60 mm, acquired matrix size 152 × 320 × 144, reconstructed resolution 0.34 × 0.34 × 1.60 mm (after two-fold interpolation in-plane by zero-filling in k-space), reconstructed matrix size 540 × 640 × 144, GRAPPA factor 2. Acquisition time: 5:25 min.

CPMG: Coronal, TR 4500 ms, TE 12 ms, number of echoes 10, echo spacing 12 ms, acquired resolution 0.68 × 0.68 × 1.7 mm inclusive of 15% slice gap, acquired matrix size 152 × 320, 34 slices, interleaved slice order, reconstructed resolution 0.34 × 0.34 × 1.7 mm (after two-fold interpolation in-plane by zero-filling in k-space, and inclusive of 15% slice gap), reconstructed matrix size 540 × 640, 34 slices, GRAPPA factor 2. Acquisition time: 11:07 min.

### Study 2

The imaging protocol included a 3D T1-weighted whole-brain MPRAGE and 2D multi-contrast turbo spin-echo (TSE).

MPRAGE: Sagittal, whole-brain, TR 2200 ms, TE 2.28 ms, TI 900 ms, flip angle 9°, FOV 220 × 220 × 179 mm, acquired resolution 0.86 × 0.86 × 0.86 mm, acquired matrix size 256 × 256 × 208. Acquisition time: 5:07 min.

Multi-contrast TSE: Coronal, TR 7500ms, number of echoes: 3, TE 9.1, 72 & 136 ms, acquired resolution 0.69 × 0.69 × 1.5 mm, reconstructed resolution 0.34 × 0.34 × 1.5 mm (after 2-fold interpolation in-plane by zero-filling in k-space, and inclusive of 15% slice gap), GRAPPA factor 2, FOV 220 × 220 × 34, acquired matrix size 270 × 320 × 58. Acquisition time: 5:09 min.

CPMG and TSE scans were not ‘whole-brain’, their coverage only extending approx. 1cm beyond anterior and posterior ends of the hippocampus. These scans were tilted such that the hippocampal body lay perpendicular to the slice acquisition plane. These scans also included the entirety of thalamus.

The two distinct methods of measuring T2 (CPMG vs TSE) will give inherently different values for T2 midpoint and heterogeneity between studies (See supplementary information). Relationships to variables such as age and cognitive score should be similar, given they are sensitive to the same tissue properties.

### Imaging analyses

All analyses were performed at CRICBristol in a Linux cluster environment. All analyses were carried out in single-subject native space.

CPMG and TSE scans were brain-extracted using FSL’s *bet2* on the first echo in the series (Smith, 2002). All extracted images were visually inspected for quality and rerun with different fractional intensity thresholds or gradient parameters where necessary. Fractional intensity threshold was typically set between 0.2-0.3. MPRAGE images were brain-extracted using *vbm8bet* (in-house script) and bias-field-corrected using FSL FAST (Zhang *et al*., 2001). T2 maps were created in MATLAB from multi-echo sequences by fitting logarithmic-space mono-exponential decay functions to each voxel series (overall summary of T2 calculation is shown in Knight *et al*. (2019)). The first echo of CPMG was always excluded. A sum-of-echoes image was created in order to have one structural image representing the entire multi-echo sequence. This image was used for segmentation.

The hippocampus and surrounding cortices were segmented using the Automatic Segmentation of Hippocampal Subfields (ASHS) software package (Yushkevich *et al*., 2015) (version: rev103, dated 12/06/2014; UPENN memory centre atlas dated 16/04/2014). CA1, CA2, CA3, dentate gyrus, subiculum and miscellaneous were combined to form a total hippocampus mask. This was overlaid onto T2 maps, giving a value of T2 for each voxel of hippocampus.

Thalamus masks were created using T1-w images as inputs to Freesurfer v6.0. After extraction from the Freesurfer segmentation image and registration to T2-space using FSL’s FLIRT, thalamus masks were then overlaid onto T2 maps, and descriptive statistics were calculated, similarly to hippocampus. These automated masking programmes have demonstrated high accuracy whilst minimising subjective rater bias, without the need for group blinding.

### Modelling T2 heterogeneity

Distribution histograms were capped at 30 ms and 200 ms, as values outside these regions are unphysiological in brain tissue at 3T. The free-to-download MATLAB function ‘fitmethis’ (de Castro, 2020) was used to fit 18 different distribution functions (See supplementary information) to left and right hemisphere ROIs individually, using maximum likelihood estimation. Akaike Information Criteria (AIC) was calculated for each distribution type. The best fitting model was determined by the lowest AIC. The most frequent best-fitting model was recorded for hippocampus and thalamus, and subsequent statistics calculated therefrom.

### Statistical analysis

Volumes, T2 metrics and cognitive scores were converted into Z-scores for each study separately and pooled, with healthy controls of each study as a reference population. Model parameters of T2 distributions were compared between groups using ANCOVA, with age as a covariate. Years of education was originally included as a covariate of all models, but did not significantly contribute to the model in any case. Reported models therefore do not correct for years of education. Homogeneity of variances was tested using Levene’s test, which was not significant for any test. Graphs show estimated marginal means from this analysis. Post-hoc pairwise comparisons were carried out using sidak correction for multiple comparisons (corrected p-values are shown as ‘psidak’). Ability of volume and T2 to predict cognitive decline was assessed using linear regression, with follow-up cognition as the dependent variable and baseline cognition and age as covariates:

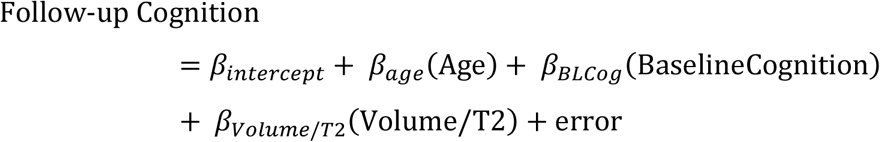

Z-scores for the latter analysis were calculated relative to each study’s MCI population only. Linear regressions were used to assess the strength of the relationship between age and T2 statistics in healthy controls. All reported p-values are two-tailed. Balance tests were not carried out on demographic for reasons detailed by Mutz *et al*. (2018).

Data handling and storage was carried out using MathWorks MATLAB 2015a (with statistics and machine learning toolbox) and Microsoft Excel 2016. Statistical analysis was performed in IBM SPSS Statistics 24. Graphs were produced using GraphPad Prism v7.

### Data availability

The data that support the findings of this study and the code used in analysis and data processing are available from the corresponding author upon request.

## Results

Demographic details for study 1 and 2 can be found separately in Table 1 & Table 2, respectively.

**Table 1.** Participant demographics for study 1 cohort. Data show mean ± standard deviation. HC = Healthy Control; MCI = Mild Cognitive Impairment; AD = Alzheimer’s disease; YOE = years of education; MoCA = Montreal Cognitive Assessment

|  | Group |  |  | Total |
| --- | --- | --- | --- | --- |
|  | HC | MCI | AD |  |
| N (male: female) | 50 (21:29) | 30 (14:16) | 10 (2:8) | 90 (37:53) |
| Age (years) | 67.7 ± 9.03 | 70.7 ± 8.55 | 77.9 ± 9.94 | 69.8 ± 9.43 |
| YOE | 15.6 ± 2.66 | 14.5 ± 2.70 | 13.1 ± 2.60 | 14.9 ± 2.76 |
| MoCA /30 | 28.0 ± 1.32 | 23.0 ± 2.83 | 16.7 ± 4.16 | 25.0 ± 4.40 |

**Table 2.** Participant demographics for study 2 cohort. Data show mean ± standard deviation. HC = Healthy Control; MCI = Mild Cognitive Impairment; YOE = years of education; ACE-III = Addenbrookes Cognitive Examination III.

|  | Group |  | Total |
| --- | --- | --- | --- |
|  | HC | MCI |  |
| N (male: female) | 47 (25:22) | 19 (13:6) | 66 (38:28) |
| Age | 71.0 ± 7.80 | 74.5 ± 9.49 | 72.0 ± 8.40 |
| YOE | 16.0 ± 3.63 | 13.7 ± 3.00 | 15.3 ± 3.59 |
| ACE-III (Total) /100 | 94.8 ± 3.20 | 80.2 ± 6.32 | 90.62 ± 7.92 |
| Attention /18 | 17.6 ± 0.64 | 15.6 ± 2.76 | 17.0 ± 1.81 |
| Memory /26 | 24.3 ± 2.05 | 16.5 ± 3.06 | 22.1 ± 4.26 |
| Fluency /14 | 12.1 ± 1.49 | 9.47 ± 2.27 | 11.3 ± 2.10 |
| Language /26 | 25.2 ± 1.04 | 23.8 ± 1.57 | 24.8 ± 1.35 |
| Visuospatial /16 | 15.6 ± 0.64 | 14.3 ± 1.59 | 15.4 ± 1.77 |

### Model fitting to describe T2 distribution characteristics

T2 distributions in the hippocampus (Figure 1) and thalamus were best described in the majority of cases by a loglogistic distribution function (as determined by the lowest AIC). Log-logistic distribution is defined as:

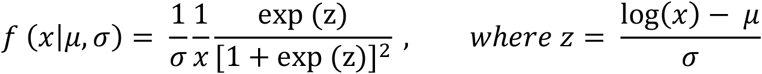

where μ and σ denote the log-median value (midpoint) and distribution shape (heterogeneity), respectively. Values for hippocampus and thalamus volume and T2 model parameters can be found in supplementary tables 1 and 2.

**Figure 1.**
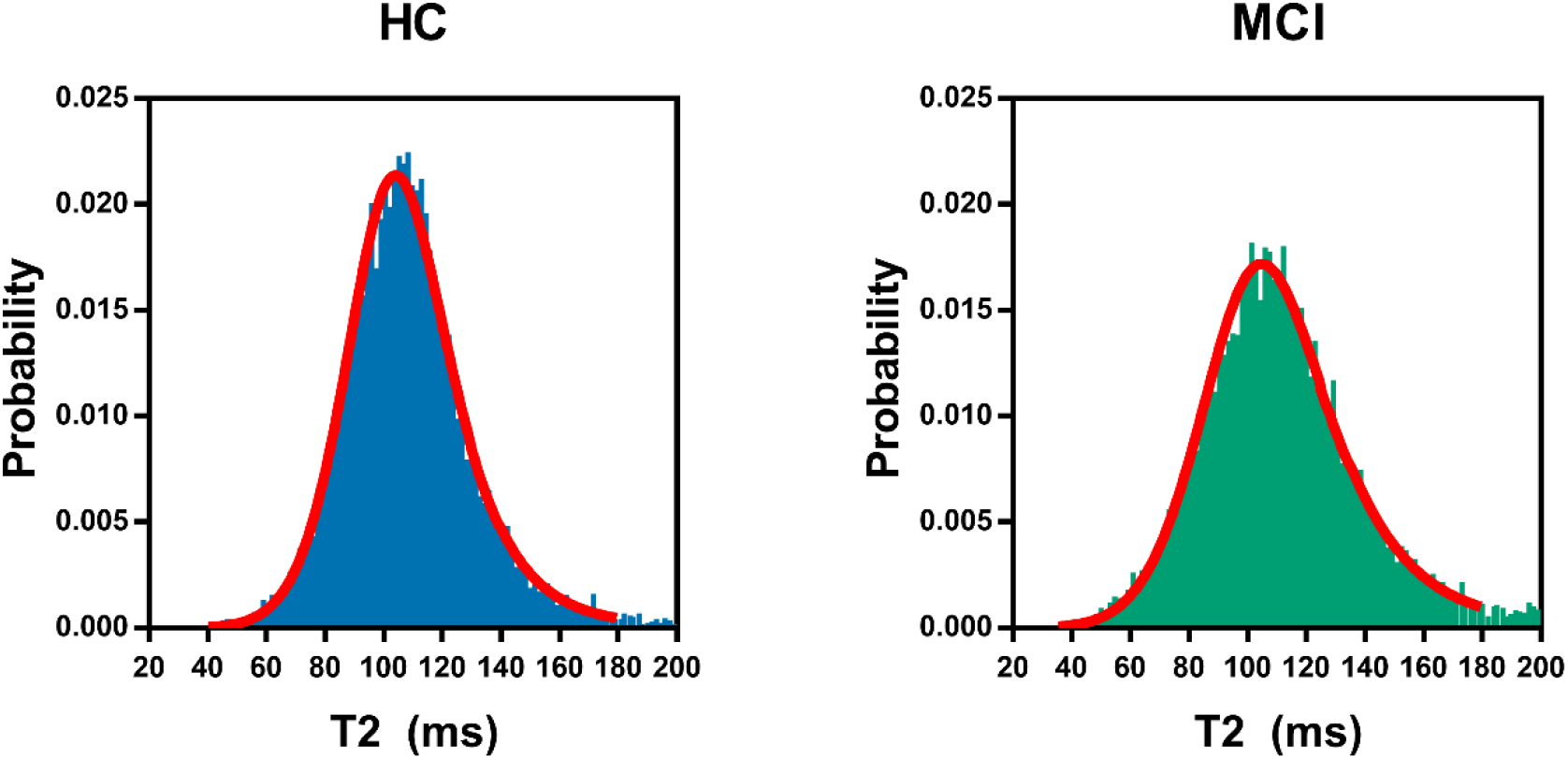
Hippocampal T2 relaxation time histograms for example participants. Left: Healthy control, 69yo female (p = 4.68, o = .112). Right: MCI, 87yo male (p = 4.71; o = .135). Left hippocampus is shown in both examples. Red lines on each graph represent loglogistic distribution curves fitted to each participant’s data.

### T2 heterogeneity, but not midpoint, differentiates healthy older adults from those with MCI

#### T2 midpoint (μ)

There was no significant difference between HC, MCI and AD groups (F(2,152)=1.61, p=.204;Figure 2A). Although T2 midpoint was higher in the AD group than other groups, this effect was not statistically significant compared to either healthy controls (p_Sidak_=.283) or the MCI group (p_Sidak_=.211).

We found a significant effect of group on T2 midpoint in the thalamus (F(2,152)=3.10, p=.048; Figure 2B). Post-hoc analyses reveal that this is driven by an increase in T2 in the AD group (HC vs AD: p_Sidak_=042; MCI vs AD: p_Sidak_=073).

**Figure 2.**
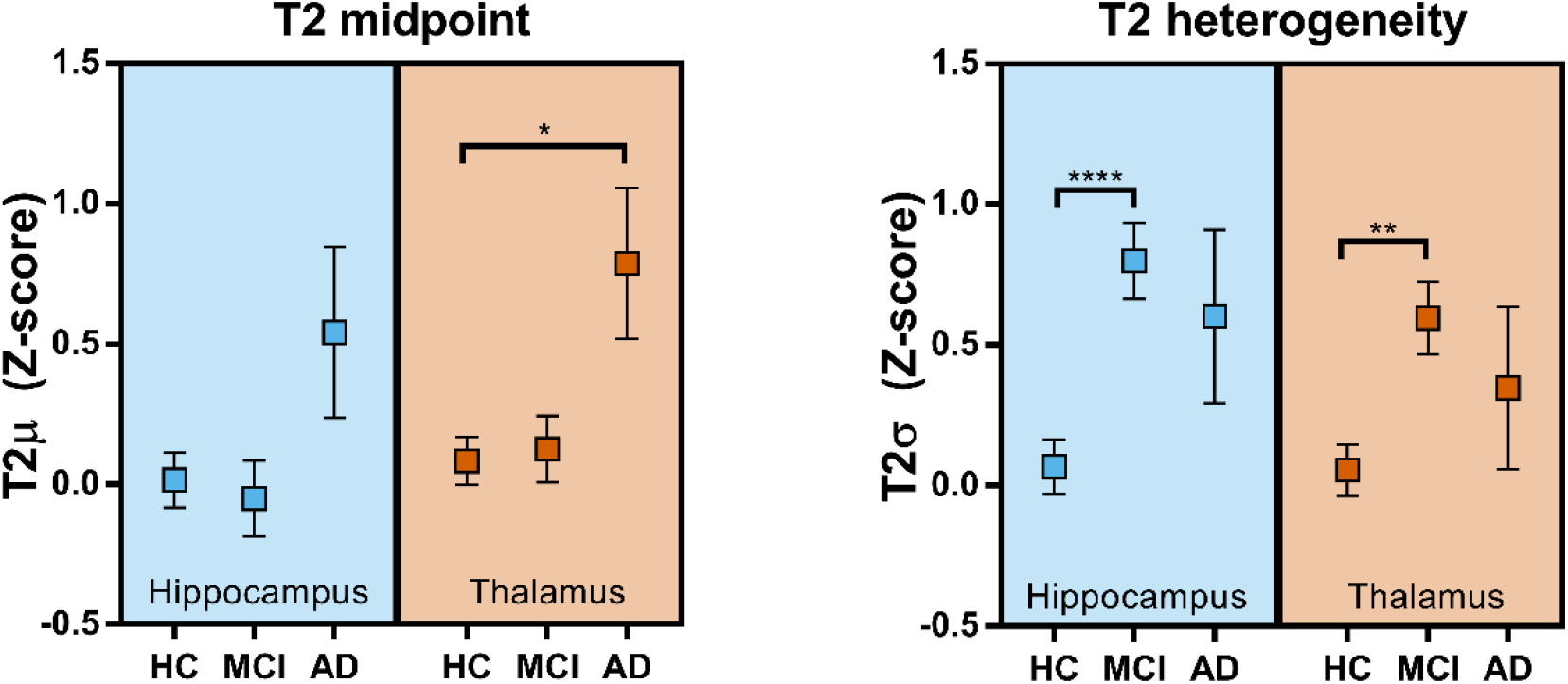
Group comparisons for T2 model descriptive parameters in hippocampus and thalamus. Values shown are estimated marginal means after correcting for the effect of age. Error bars show marginal means ±standard error. Asterisks represent Sidak pairwise comparisons p-values (*p<.05; **p<.01; ****p<.0001).

#### T2 heterogeneity (σ)

There was a significant effect of group on hippocampal T2 heterogeneity (F(2,152)=9.76, p=.0001). Pairwise comparisons reveal a significantly wider distribution in the MCI group compared to healthy controls (p_Sidak_<.0001). There was no significant further change from MCI to AD (p_Sidak_=913).

There was a significant difference in thalamic T2σ between groups (F(2,152)=5.90, p=.003). Post-hoc pairwise comparisons reveal a significantly increased T2σ in the MCI group compared to HC_s_ (p_Sidak_=.002).

### T2 heterogeneity predicts cognitive decline in Mild Cognitive Impairment

Hippocampal T2 heterogeneity significantly predicted follow-up cognitive score, after accounting for baseline cognitive score and age (R^2^=.387, F(3,16)=3.37, p=.045; Figure 3A). T2 heterogeneity was the sole significant individual predictor in this model (β_T2σ_=-.601, pT2σ=.018). Cognitive change over time was not predicted by this method by either hippocampal T2μ (R^2^=.241, F(3,16)=1.69, p=.209; β_T2μ_=-.377, p_T2μ_=132; Figure 3B) or hippocampal volume (R^2^=.177, F(3,16)=1.15, p=.361; βvol=.263, pvol=.315; Figure 3C).

**Figure 3.**
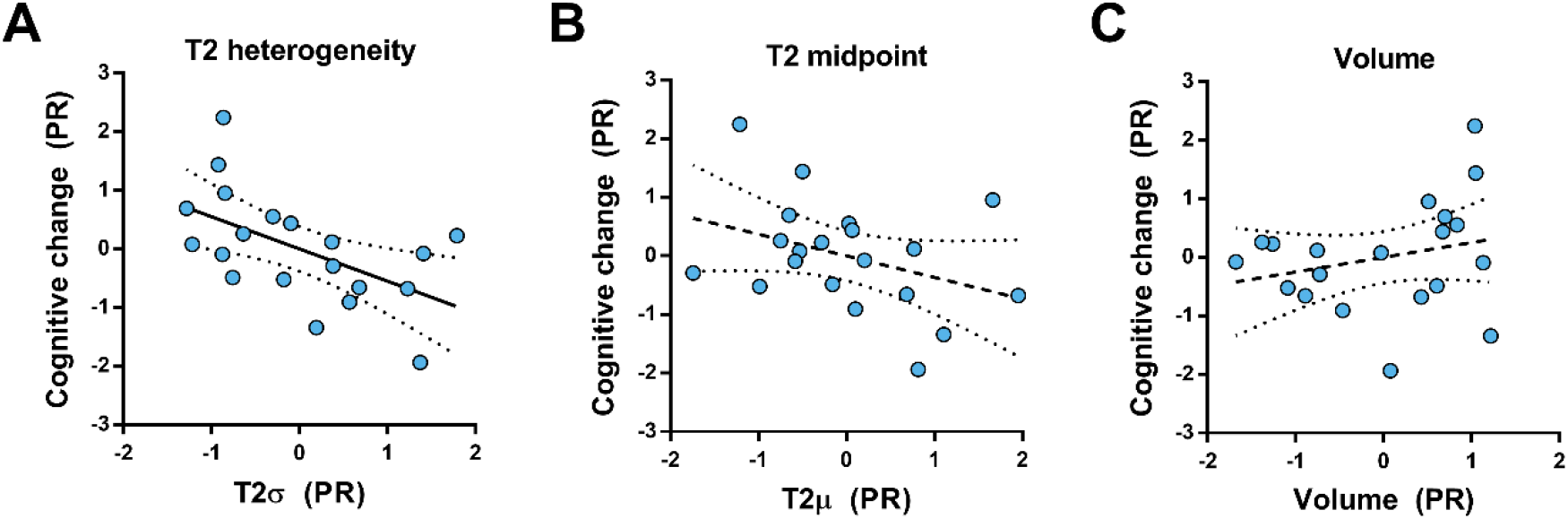
The ability of hippocampal metrics to predict cognitive change over one year. Data shown are partial residual (PR) plots for hippocampal structural measures predicting follow-up cognitive score, correcting for age and baseline cognition. Y-axis shows standardised residuals from linear regression of age and baseline cognitive score predicting follow-up cognitive score. X-axes also show standardised residuals with the same predictors, predicting hippocampal volume (A: volume normalised to ICV), hippocampal T2 midpoint (B) or hippocampal heterogeneity (C). Solid black lines represent linear regression slopes with p<.05. Dotted lines represent those with p>.05. Regression lines are shown with ± 95% confidence intervals.

### T2 relaxometry in healthy ageing

#### T2 midpoint (μ)

There was no statistically significant relationship between age and hippocampal T2 midpoint (T2μ, R^2^=.012, p=.289, n=97; Figure 4A). In the thalamus, age was a strong positive predictor of T2μ (R^2^=.320, p<.0001, n=97; Figure 4B).

**Figure 4.**
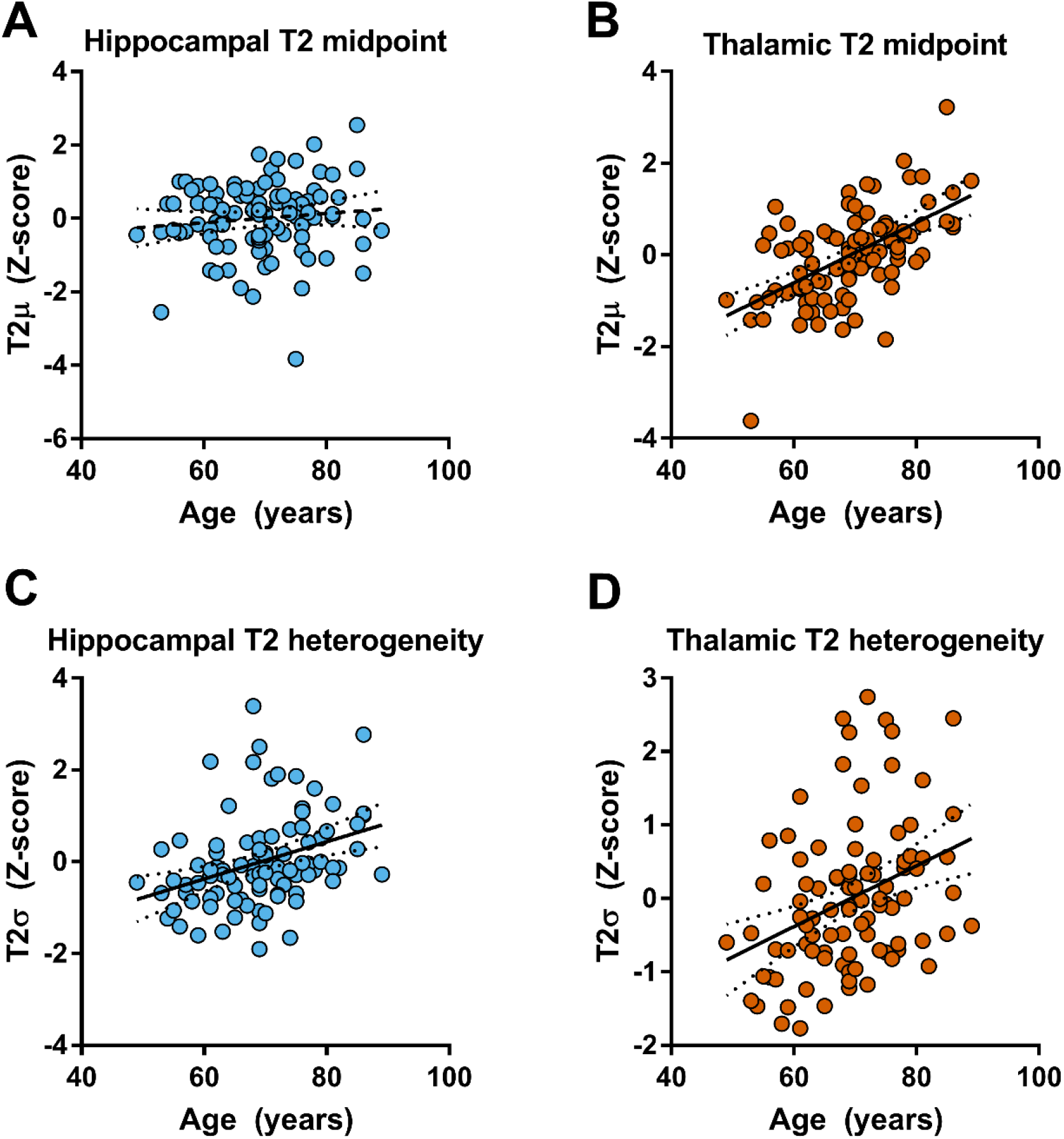
Linear regressions for age predicting T2 model descriptive parameters in hippocampus and thalamus. Regressions shown are between age and hippocampal T2p (A), thalamic T2p (B), hippocampal T2o (C) and thalamic T2o (D). Solid black lines represent linear regression slopes with p<.05. Dashed lines represent those with p>.05. Regression lines are shown with ± 95% confidence intervals.

#### T2 heterogeneity (σ)

Age was a significant positive predictor of T2 heterogeneity in the hippocampus (T2σ, R^2^=.122, p=.0004; Figure 4C) and thalamus (R^2^=.127, p=.0003; Figure 4D).

## Discussion

We show that the width of the distribution of T2 in the hippocampus and the thalamus differentiates healthy older adults from those with Mild Cognitive Impairment, while the T2 midpoint does not. Heterogeneity of T2 may therefore be an early marker of structural integrity in Alzheimer’s disease. Although ageing affects T2, even after controlling for age, T2 heterogeneity predicted decline whereas hippocampal volume and T2 midpoint did not.

Based on the presented T2 relaxometry data, we propose the following model in Figure 5 where healthy ageing is characterised by a relative dominance of factors that increase T2 over factors that decrease T2, particularly in the thalamus. Incipient Alzheimer’s disease is characterised by additional factors that decrease T2, balancing out the effects of T2-increasing factors on T2 midpoint to some extent. This leads to an increasing width of the distribution of T2 without necessarily changing the midpoint in prodromal Alzheimer’s disease (MCI). In later stages of disease, after a diagnosis of Alzheimer’s disease, factors that increase T2 predominate. This model explains these data and ties together previous seemingly conflicting literature such as the discrepancy between human and animal literature of T2 changes due to Alzheimer’s disease.

**Figure 5.**
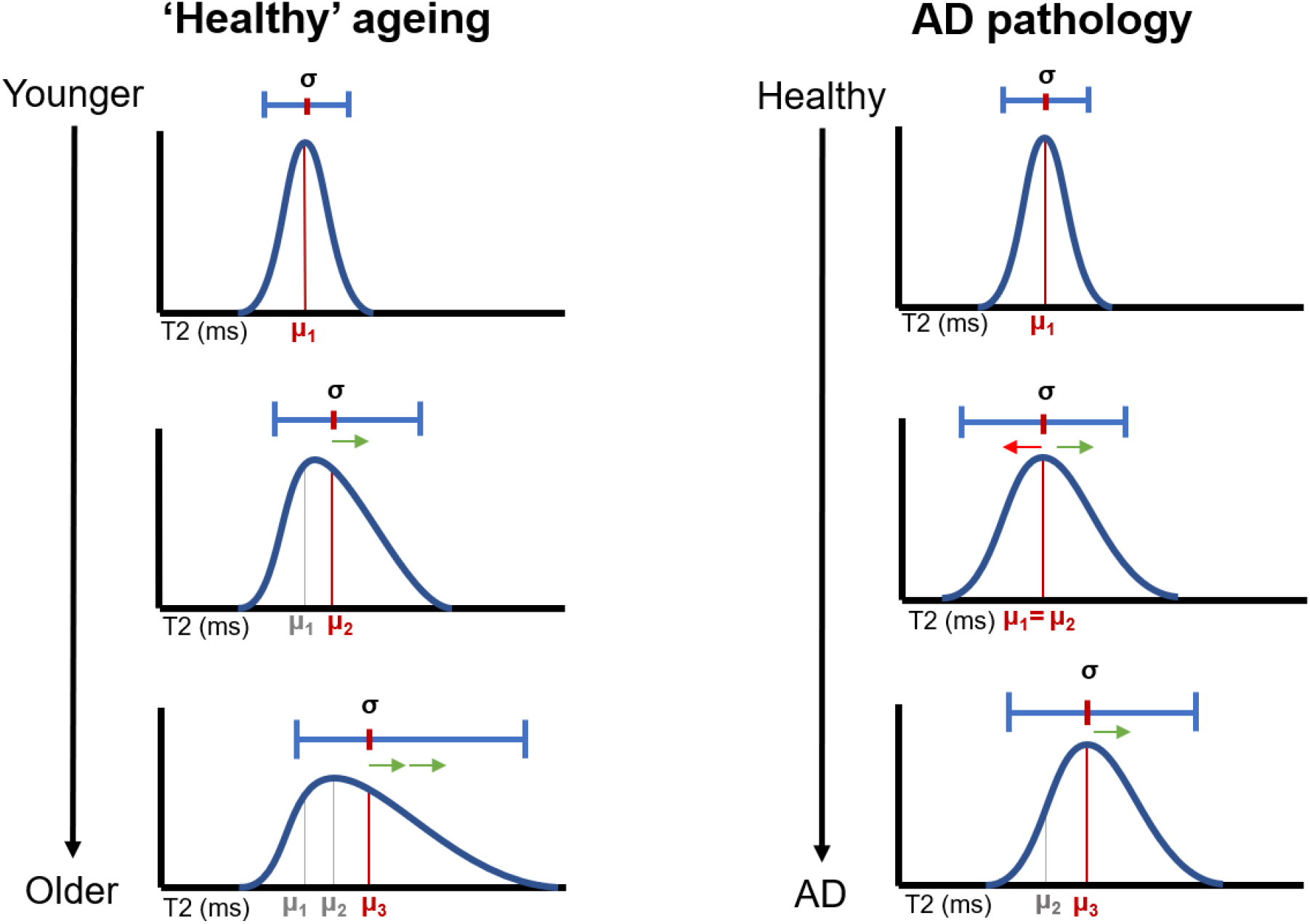
Schematic diagram of T2 distribution profiles in ageing and Alzheimer’s disease. Midpoint values for each hypothetical distribution are represented by orange bars and “μ” markers on each × axis. Green and red arrows represent factors that increase or decrease T2, respectively. The number of arrows represents the relative dominance of each effect. In summary the model suggests that factors that increase T2 are present in both healthy ageing and Alzheimer’s pathology, however factors that decrease T2 are much more specific to, or dominant in Alzheimer’s disease. Early Alzheimer’s disease pathology is characterised by an increase the distribution without an increase in the mean.

### T2-prolonging factors are dominant in healthy ageing and later stage Alzheimer’s disease

The primary causes of variability in T2 are content and mobility of water. T2 increases as water mobility increases (Kamman *et al*., 1988). The amount of free water in a region can be partially attributed to the inverse of the compartmentalisation of the water, as is caused by cell membrane disruption. As cells die, whether due to normal ageing processes (Golomb *et al*., 1993; Pol *et al*., 2006) or Alzheimer’s pathology, cell membranes become damaged, thereby increasing the amount of free water within a tissue (Besson *et al*., 1992). The breakdown of myelinated structures also causes T2 to increase in white matter and can be caused by both ageing and Alzheimer’s disease (Paus *et al*., 2001; Bartzokis *et al*., 2004; Bartzokis *et al*., 2006; Bartzokis, 2011; Alonso-Ortiz *et al*., 2015; Knight *et al*., 2016). This leads to an increase in T2 both in healthy ageing and in Alzheimer’s disease, even in early stages, as a result of microstructural damage. In support of this, this study shows a significantly longer T2 in the thalamus of older people and Alzheimer’s disease patients compared to healthy controls.

Additionally, we show that T2 heterogeneity is predicted by age in both thalamus and hippocampus, an effect that would be expected from uneven increases in T2 across the region.

However, T2 does not appear to increase significantly in the hippocampus either with age or disease progression, except perhaps at later stages of the disease course, after a diagnosis of Alzheimer’s disease. One explanation is that the increase in T2 is balanced out in the hippocampus by T2-shortening factors that are present even prior to MCI diagnosis. This is discussed further in the following sections.

### T2-shortening factors are indicative of Alzheimer’s pathology

An established biomarker of Alzheimer’s disease is the build-up of large extracellular oligomers of Aβ and intracellular NFTs. In early stages of the disease these plaques and tangles are disparate and unevenly spread throughout the brain (Braak and Braak, 1991; Braak and Braak, 1995; Thal *et al*., 2002). The hippocampus is one of the first regions to start expressing these hallmarks (Thal *et al*., 2002). The thalamus is also affected very early on, with potential signs showing in Braak stage 1 (Braak and Braak, 1991). The presence of plaques or NFTs will cause T2 to decrease locally due to an increased macromolecule-to-water ratio and the restriction of water motility in the extracellular space, even before any neurotoxic damage that may be caused by these structures. Even before the build-up of plaques, T2 has been shown to be decreased in the hippocampus in animal models (El Tannir El Tayara *et al*., 2006; Falangola *et al*., 2007). T2 can also be shortened by paramagnetic materials such as iron (Hardy *et al*., 2005; El Tannir El Tayara *et al*., 2006; Jara *et al*., 2006; Meadowcroft *et al*., 2015), which is associated with neurodegenerative disorders like Alzheimer’s disease and Parkinson’s disease (Smith *et al*., 2010; Castellani *et al*.,2012; Ward *et al*., 2014).

Participants with incipient Alzheimer’s disease might therefore be expected to have decreased T2 in the hippocampus due to the overexpression of Aβ and NFTs, and accumulation of iron. This would balance out the T2-prolonging factors discussed previously. In support of this, this study shows a substantial increase in distribution width of T2 in both hippocampus and thalamus, after correcting for age, in people with MCI compared to healthy controls. Furthermore, the study also shows no increase of T2μ in MCI compared to controls, a result to be expected given counteracting factors increasing and decreasing T2.

The described model is further supported by a previous study by Su *et al*. (2016). The crosssectional results of their study revealed that Alzheimer’s disease patients had significantly reduced T2 compared to healthy controls. However, longitudinally, T2 in Alzheimer’s disease patients was seen to increase. The currently presented model explains these results in terms of a shift in the dominance of factors that increase or decrease T2 throughout the progression of Alzheimer’s disease. Alzheimer’s disease-specific macromolecular pathological hallmarks cause T2 to decrease in the first instance, which later causes physical damage to the structure, causing T2 to increase as the disease progresses (Figure 6), as is seen in the majority of studies on T2 in Alzheimer’s disease (Kirsch *et al*., 1992; Laakso *et al*., 1996; Pitkänen *et al*., 1997; Wang *et al*., 2004; Dawe *et al*., 2014).

There is, of course, considerable debate as to the role of plaques in Alzheimer’s disease pathology (for a review see (Morris *et al*., 2014)) and some question as to the disease-specificity of iron accumulation (Castellani *et al*., 2012; Ward *et al*., 2014). Indeed, plaques have been found in the brains of many people without any other sign of Alzheimer’s disease, particularly in the hippocampus (Arriagada *et al*., 1992; Davis *et al*., 1999). Oligomeric Aβ, however, could still have T2-shortening effects in the brains of people with early Alzheimer’s disease. The presence of some T2-shortening factors even in those with no Alzheimer’s disease-specific pathology could explain the lack of correlation between hippocampal T2 midpoint and age in healthy control participants. An example of this could be caused by iron in microglia which are recruited in response to inflammation. Although inflammation is a factor in Alzheimer’s disease, it could also be present in response to comorbidities like cardiovascular disease. Conversely, cardiovascular disease could also reduce blood flow to the brain, potentially reducing the iron and causing T2 to increase. Unfortunately, amyloid and iron status of the current studies’ participants were not available, so the higher number of voxels with low T2 cannot be directly attributed to either factor. Future work could aim to colocalise areas of low T2 with Aβ, for example with positron emission tomography (PET), and brain iron levels by measuring field dependent relaxation rate increase (FDRI) as was done by Raven *et al*. (2013).

Despite some presence of Aβ and iron in healthy ageing, various studies suggest that the two factors combine in Alzheimer’s pathology, leading to the much greater T2-shortening effects seen in Alzheimer’s disease. A study by El Tannir El Tayara *et al*. (2006) showed that T2 in the hippocampus (specifically in the subiculum), was decreased in a mouse model of Alzheimer’s disease that produced amyloid deposits (APP/PS1), compared to another model that does not form such deposits (PS1). The authors attribute this, at least in part, to the colocalization of amyloid and iron. Such histological colocalization has also been reported by Falangola *et al*. (2005). Excess iron can not only contribute to oxidative stress in and of itself, but can also contribute to Aβ and NFT misfolding (Sayre *et al*., 2000), thereby exacerbating Alzheimer’s pathology. Various studies are supportive of the idea that a combination of iron and Aβ cause significant T2 shortening (Bartzokis *et al*., 1994; Savory *et al*., 2002; Helpern *et al*., 2004; Falangola *et al*., 2005; El Tannir El Tayara *et al*., 2006; Falangola *et al*., 2007; House *et al*., 2008; Qin *et al*., 2011; Teipel *et al*., 2011).

### Discrepancies between studies of T2 in humans and transgenic rodent models

The proposed model can also begin to explain the discrepancy between studies of T2 in rodent models of Alzheimer’s disease and human cases. As summarised in a recent review (Tang *et al*., 2018), the majority of studies to date looking at T2 alterations in rodent models of Alzheimer’s disease observe decreased T2 in the hippocampus. However, in human literature the picture is much more varied, with most studies reporting a T2 increase (Kirsch *et al*., 1992; Laakso *et al*., 1996; Pitkanen *et al*., 1996; Wang *et al*., 2004; Raven *et al*., 2013), some reporting no change at all (or changes in just one hemisphere) (Laakso *et al*., 1996; Campeau *et al*., 1997), and some reporting a decline (House *et al*., 2006; Luo *et al*., 2013). Considering the currently presented model, this suggests that the animal models of Alzheimer’s disease used to test this effect have an over-dominance of T2-shortening factors. Given that these transgenic models are overwhelmingly based on mutations within amyloid cascade pathways, it is reasonable to conclude that these models may lead to a more dominant effect of Aβ on T2 in these models than is the case in “human” Alzheimer’s disease. Indeed, many transgenic rodent models express an abundance of Aβ plaques but show little or no neurodegeneration (Philipson *et al*., 2010), and therefore have fewer regions of increased T2. Furthermore, most people with late-onset Alzheimer’s disease do not have mutations in any of the primary genes modified in rodent models. Many of these transgenic mouse models are models of amyloidopathy rather than models of Alzheimer’s disease; they do not necessarily reflect the heterogeneity seen in Alzheimer’s disease in humans.

### Differences between T2 profiles in hippocampus vs thalamus

The overall pattern of results is very similar between hippocampus and thalamus. However, a primary difference is that age is a strong predictor of T2 midpoint only in the thalamus. By only looking at T2 midpoint, it is easy to conclude that the hippocampus is resistant to age and the thalamus is sensitive, as is concluded by Kirsch *et al*. (1992). However, when looking at the distribution of T2, it is clear that the hippocampus is not unaffected by age, but rather, factors that increase and decrease T2 are more evenly balanced than in the thalamus. Given that loci of amyloid deposition throughout the brains of both healthy and Alzheimer’s disease patients is similar (Arriagada *et al*., 1992; Thal *et al*., 2002), Aβ is more likely to be found in the MTL than in the thalamus, even in individuals with no Alzheimer’s disease symptoms. This supports the idea that even in “healthy” hippocampi, some Aβ will be causing T2-shortening coincident with T2-lengthening factors dominant in aging, the latter effects only becoming dominant in much older people.

The effect size of group on T2 heterogeneity was stronger for hippocampal T2 than thalamic T2. These results reflect previous literature on the severity of histopathological changes in either region (Braak *et al*., 1996). Here, Braak & Braak report that the thalamus, unlike the hippocampus, is relatively resistant to atrophy in Alzheimer’s disease until relatively late stages, despite early deposition of Aβ. As a result, the increase in T2 heterogeneity may be an early indicator of imminent neuronal loss. An alternate explanation is that early changes in T2 only occur in a small portion of the thalamus. Future work will explore individual hemisphere effects and subfields of both the thalamus and hippocampus, though this is beyond the scope of this paper.

Both of these differences support the use of T2 heterogeneity over T2 midpoint as a measure of tissue integrity.

### Potential clinical utility of hippocampal T2 relaxometry

Understanding T2 dynamics in preclinical Alzheimer’s disease and healthy ageing offers the potential for great clinical benefit. If Alzheimer’s pathology can be detected using MRI prior to the onset of hippocampal atrophy, significant change in cognition, or loss of daily independence patients may receive treatment much earlier – at a stage where neurodegenerative damage is preventable or even reversable.

Hippocampal volume is the one of the most widely studied and effective predictors of cognitive decline (for a review see de Flores *et al*. (2015)). However, rather than measuring pathology itself, volumetry measures tissue atrophy, a consequence of pathology. T2 increases also measure consequences of pathology, in the form of increased regional CSF, oedema or cell membrane breakdown, however it is a more sensitive measure, and may indicate subtle damage before macroscopic atrophy is visible. Furthermore, T2 decreases may measure key features of Alzheimer’s pathology itself, such as iron, Aβ and NFT deposition that can occur before hippocampal shrinkage (Jack *et al*., 2008). Measuring T2 heterogeneity allows these opposing factors to be considered, as they may indicate slightly damaged tissue that has the potential for therapeutic rescue. Measuring T2 distribution width compared to age-corrected normative data may be indicative of physical damage beyond what should be expected for a given age. These markers may compliment or even surpass volumetry in predicting future cognitive decline. As neuroimaging, often MRI, is part of routine clinical screening processes for neurological disease, this method is highly practical and easily translatable.

## Limitations

With the exception of some of those who have actually received a diagnosis of Alzheimer’s disease in study 1, the amyloid status of these participants is unknown. Amyloid is one of the most commonly used biomarkers to increase certainty of the presence of Alzheimer’s disease pathology. Those who present with mild cognitive impairment often are only classified as MCI based on presentation of cognitive symptoms. Such memory impairment could be caused by any number of factors other than Alzheimer’s pathology, including other dementias, stroke, pharmaceutical side effects and sleep problems to name a few. This being said, around half of MCI patients do later receive diagnoses of Alzheimer’s disease (Fischer *et al*., 2007), so it is likely that impairment in these cohorts would be attributable to Alzheimer’s disease.

Secondly, although we present results in a large sample of healthy older controls and people with MCI, we are limited by our small sample of Alzheimer’s disease patients. This is primarily because they were only recruited as a part of study 1. This limits the statistical significance of some of the effects that we describe, and therefore conclusions from this group are slightly tentative. This is acknowledged throughout interpretation of these results, which we expect to be reproducible with a larger sample size.

Finally, the only regions studied here, hippocampus and thalamus, are both regions known to be affected by Alzheimer’s pathology at early stages. Future studies would benefit from exploring T2 dynamics in other brain regions, including those that are not directly implicated in early Alzheimer’s disease. This is not possible with existing data for either study 1 or study 2, as the multi-echo T2 scans acquired do not cover the whole brain.

## Conclusions

In this paper, we show that T2 heterogeneity is a good measure of microstructural integrity of brain tissue. We propose a model (Figure 5) that suggests factors that increase T2 are indicative of microstructural damage but are not necessarily specific signs of Alzheimer’s pathology. Rather, factors that decrease T2 are much more specific to and dominant in Alzheimer’s pathology and may occur in the earliest stages of disease (Figure 6). These two opposing forces act to balance out the mean in prodromal Alzheimer’s disease, causing varied results in the human literature. The model makes specific and testable predictions about the temporal dynamics of T2 alterations throughout ageing and prodromal Alzheimer’s disease. It also highlights potential early indicators of Alzheimer’s disease, allowing Alzheimer’s disease-related cognitive decline to be distinguished from that seen in healthy ageing. We show that T2 heterogeneity surpasses midpoint T2 and the more established measure of volumetry in predicting cognitive decline in those with MCI.

**Figure 6.**
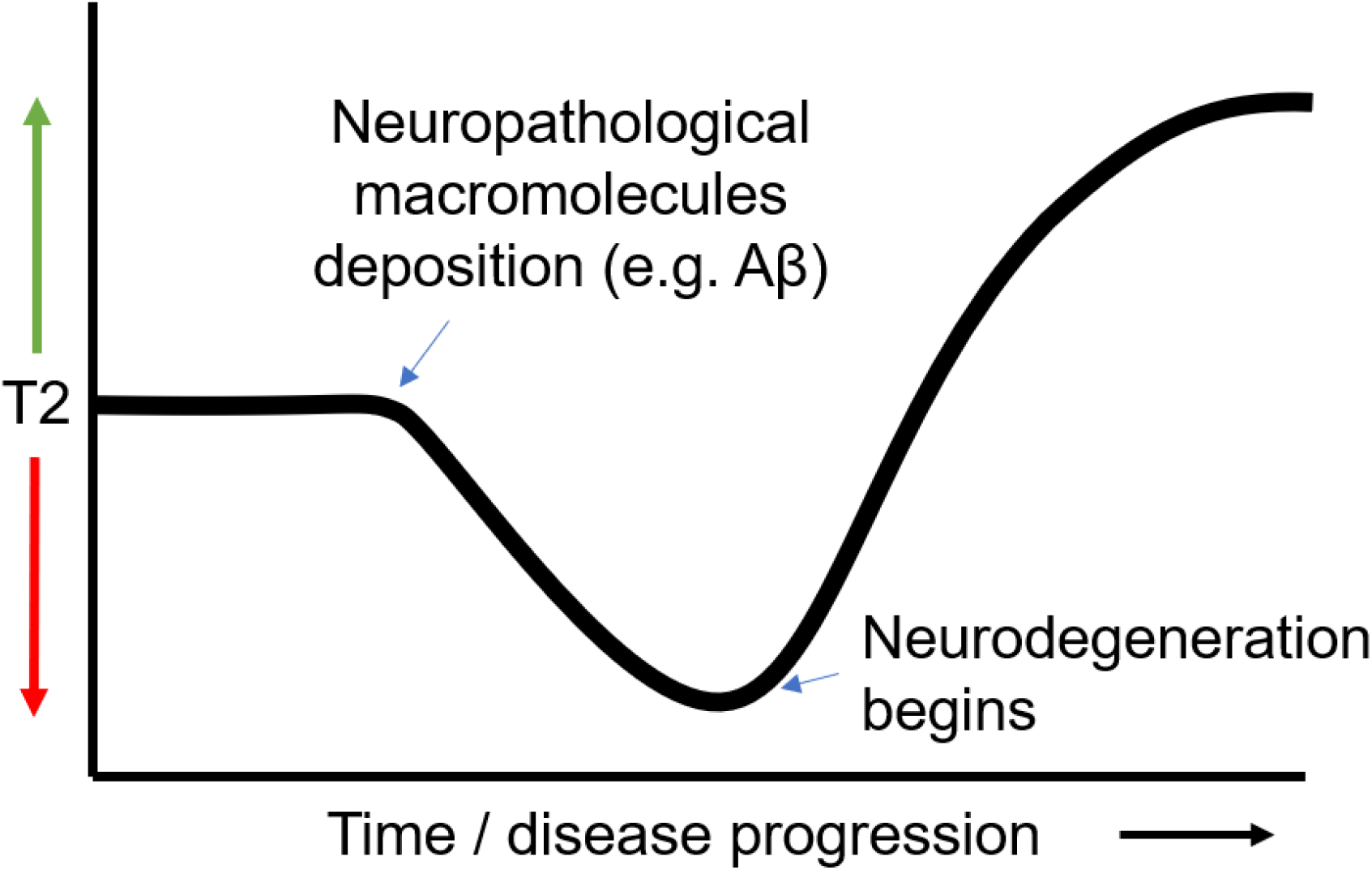
Theoretical model of T2 dynamics in a single given voxel in the brain throughout the course of Alzheimer’s disease. A given region in brain of someone with incipient Alzheimer’s disease would consist of many voxels at different stages of this curve, depending on the degree of Alzheimer’s pathology in a given location. This heterogeneity is what will cause the average or midpoint T2 to remain relatively static, and the distribution width to increase, until very late stages when all voxels reach the ‘high T2’ state.

This study represents one of the first studies into the utility of the T2 heterogeneity within the brain in the context of Alzheimer’s disease, and the first to show its utility in predicting cognitive decline in groups at-risk for Alzheimer’s disease.

## Data Availability

The data that support the findings of this study are available from corresponding authors upon request. 

## Acknowledgments

The authors wish to thank Join Dementia Research and the Avon & Wiltshire Mental Health Partnership for their assistance with participant recruitment. We also wish to thank those who have helped collect data for the projects (Emma Hadley, Ellen Gaaikema, Lucy Adams, Candida Stainer, Ben Kershaw & Bryony McCann) and all the volunteers who gave up their time to take part in our studies.

## Abbreviations

Ap: Beta-amyloid
ACE-III: Addenbrooke’s cognitive examination version 3
AD: Alzheimer’s disease
AIC: Akaike Information Criteria
HC: Healthy control
MCI: Mild cognitive impairment
MoCA: Montreal cognitive assessment
MPRAGE: Magnetization-prepared rapid gradient echo
T2μ: T2 midpoint model parameter
T2σ: T2 distribution width model parameter (heterogeneity)
TSE: Turbo spin-echo
YOE: Years of education

